# The neural basis of prosocial effort-based decision making in older adults at risk for Alzheimer’s disease

**DOI:** 10.64898/2026.01.10.26343857

**Authors:** Caitlin S. Walker, Garance Barnoin, Mitchell Bennett, Colleen Hughes, Jennifer Tremblay-Mercier, R. Nathan Spreng, Judes Poirier, Laurel S. Morris, the PREVENT-AD Research Group, Sylvia Villeneuve, Maiya R. Geddes

## Abstract

**Background:** Alzheimer’s disease is associated with impairments in decision making that undermine autonomy, health behaviors, and quality of life. Effort-based decision making, the process of weighing reward value against effort costs, is particularly disrupted in aging and Alzheimer’s disease. However, aging is also characterized by a shift toward socioemotional and prosocial goals, which may preserve motivation and effortful behavior. Understanding prosocial effort-based decision making and its neural substrates in individuals at risk for AD may reveal early alterations in decision making and neural circuits that support healthy aging.

**Methods:** Fifty-two older adults from the PREVENT-AD cohort (mean age = 68.48, 38 females, 18 *APOE4* carriers) completed an effort-based decision-making task comparing willingness to exert effort for rewards obtained for oneself or for charity. Decision response, response time, and vigor were analyzed using mixed-effects models. Reward–effort relationships were modeled using psychometric functions and compared using random-effects Bayesian model comparison, with parameter estimates from the winning model compared between conditions and between *APOE4* carriers and non-carriers. Seed-to-voxel resting-state functional connectivity from the ventromedial prefrontal cortex and anterior cingulate cortex examined neural substrates of prosocial effort-based decision making, and ROI-to-ROI connectivity between regions of a frontostriatal reward network was compared between *APOE4* carriers and non-carriers.

**Results:** Participants showed increased acceptance of effort for prosocial compared to self-oriented rewards. The relationship between reward and effort in both conditions was best captured by a sigmoid function, with lower motivation for self-oriented compared to prosocial rewards. Across all participants, higher motivation for prosocial compared to self-oriented rewards was associated with resting-state functional connectivity to the ventromedial prefrontal cortex and anterior cingulate cortex. *APOE4* carriers showed lower overall motivation than non-carriers, but higher vigor when exerting effort for prosocial than self-oriented rewards, along with reduced nucleus accumbens–dorsal anterior cingulate connectivity that was associated with lower motivation for effort.

**Conclusions:** Prosocial incentives may be an effective strategy for motivating effortful behavior in older adults at risk for Alzheimer’s disease. Although *APOE4* carriers show greater aversion to accepting effort for both reward types, they exhibit heightened vigor when working for prosocial compared to self-oriented rewards. Leveraging prosocial motivation and its underlying neural circuitry may therefore represent a promising strategy to sustain goal-directed behavior and decision making, promote physical and cognitive activity, and support emotional and brain health in at-risk aging.

## Introduction

Individuals with Alzheimer’s disease (AD) often show impaired decision-making capacity, which is associated with reduced autonomy in activities of daily living, diminished ability to make informed health-related decisions, increased financial exploitation, and heightened caregiver burden.^1–3^ Effort-based decision making is the process by which individuals evaluate whether a reward’s value justifies the effort required to obtain it.^4,5^ Compared to younger adults, cognitively unimpaired older adults tend to avoid high-effort options and show heightened sensitivity to cognitive and physical effort costs.^6,7^ Individuals with mild cognitive impairment (MCI) and probable AD with elevated apathy demonstrate reduced sensitivity to effort costs characterized by a bias toward rejecting effortful options, potentially due to declines in executive functioning.^4,5^ Disruptions in effort-based decision making may hinder engagement in modifiable behaviors that protect physical and cognitive health, including physical activity and social and cognitive engagement.^8–10^

Despite these age-related changes in effort-based decision making, aging is also characterized by shifts in social motivational priorities. Socioemotional Selectivity Theory posits that as perceived time horizons shrink with age, individuals increasingly prioritize emotionally meaningful social goals over information acquisition.^11,12^ Consistent with this framework, older adults show enhanced positive affect and a greater focus on maintaining close social relationships relative to younger adults.^13–16^ Moreover, decision-making abilities are often preserved in emotionally salient and interpersonal contexts in older adulthood.^17,18^ Compared to younger adults, older adults also engage in more prosocial behaviors, such as volunteering and charitable giving, and display a greater willingness to expend effort that benefits others.^9,19–21^ Enhanced prosocial behavior is particularly relevant in preclinical AD, as socioemotional processes appear preserved or even enhanced in early disease stages.^22^ Moreover, prosocial behavior in later life is associated with greater purpose in life, improved physical and cognitive health, and reduced loneliness and depression.^23–25^

At the neural level, prosocial behavior consistently implicates the ventromedial prefrontal cortex (vmPFC) and the anterior cingulate cortex (ACC). The vmPFC integrates reward value and effort costs to compute subjective value, particularly in socioemotional contexts.^26–28^ In younger adults, increased vmPFC activity during a compassion exercise predicted subsequent reductions in sedentary behavior, highlighting its role at the intersection of interpersonal experience and physical effort.^29^ VmPFC activity has also been shown to predict decisions to purchase social products that benefit others rather than commercial products.^30^ Moreover, individuals with vmPFC lesions or atrophy, including those with frontotemporal dementia, exhibit reduced generosity, trust, and empathy, and discount rewards by effort more steeply when rewards benefit others.^31–34^ In addition to the vmPFC, neurophysiological recordings in non-human primates show that ACC neurons selectively encode rewards allocated to others and track others’ identities in social contexts.^35,36^ Lesions to the ACC reduce prosocial learning and behavior in both animals and humans,^33,37^ and human fMRI studies show heightened ACC activity when individuals exert effort to obtain rewards for others rather than themselves.^38,39^ Together, these findings suggest that the vmPFC and ACC are core components of the neural architecture supporting prosocial effort-based decision making.

The apolipoprotein E ε4 (*APOE4*) allele is the strongest known genetic risk factor for late-onset AD.^40,41^ *APOE4* carrier status has been associated with age-related cognitive decline, including impairments in executive function and goal-directed behavior.^42–44^ Notably, *APOE4* carriers relative to non-carriers often show heightened cognitive reserve and a reduced risk of future cognitive decline from engaging in effortful healthy lifestyle behaviors, including physical activity and social engagement.^45–49^ At the neural level, the *APOE4* allele contributes to neural vulnerability through mechanisms such as neuroinflammation, hyperexcitability, impaired lipid regulation, mitochondrial dysfunction, and amyloid dysregulation, which may disrupt neural circuits supporting motivated behavior.^50^

The dorsal anterior cingulate cortex (dACC) and vmPFC are key cortical nodes within frontostriatal reward circuits that integrate motivational, cognitive, and motor processes during decision making.^51,52^ These regions receive dopaminergic input from the ventral tegmental area (VTA) and nucleus accumbens (NAcc) and project to prefrontal regions to guide effortful behavior.^52^ Animal models demonstrate that ACC lesions or ACC–NAcc disconnections reduce willingness to exert effort,^53,54^ whereas human neuromodulation studies show that dACC stimulation evokes strong perseverance to overcome challenges.^55,56^ In aging, preserved dACC structure and connectivity are associated with exceptional perseverance and tenacity to achieve goals seen in “superagers”.^57^ However, cognitively unimpaired *APOE4* carriers exhibit early structural and functional alterations to regions within the reward network, including structural vulnerability of the VTA, elevated amyloid and tau deposition in the ACC and vmPFC, and reduced functional connectivity, cortical thickness, cerebral blood flow, and glucose metabolism of the NAcc, ACC, and vmPFC,^58–64^ potentially impacting frontostriatal reward circuits supporting effort-based decision making. Resting-state fMRI provides an ideal approach for this question because it captures intrinsic functional organization of brain networks and is sensitive at detecting early brain changes associated with AD pathology prior to structural atrophy.^65,66^

The primary aim of the present study was to compare effort-based decision making for prosocial versus self-oriented rewards in older adults at risk for AD and to identify neural substrates associated with prosocial effort-based decision making. We hypothesized that, across participants, at-risk older adults would exert greater motivation (i.e., higher acceptance of effort, lower response times, higher vigor, higher reward sensitivity, and lower bias away from effort) for prosocial relative to self-oriented rewards, and that prosocial effort-based decision making would be associated with greater resting-state functional connectivity (rsFC) to the vmPFC and ACC, regions implicated in social and motivational valuation. Given evidence that the *APOE4* allele is associated with alterations in regions of the frontostriatal reward network and reduced goal-directed behavior, it was further hypothesized that *APOE4* carriers would show overall impairments in effort-based decision making and altered rsFC between key reward regions, including the VTA, NAcc, dACC, and vmPFC, relative to non-carriers. Exploratory analyses additionally examined whether *APOE4* status differentially modulated prosocial versus self-oriented effort-based decision making.

## Materials and methods

### Participants

Participants were 52 older adults from the Presymptomatic Evaluation of Experimental or Novel Treatments for Alzheimer’s Disease (PREVENT-AD) cohort at McGill University.^67^ The sample size was calculated based on a paired *t*-test model with a two-tailed alpha of *p* < .05, a moderate effect size of Cohen’s *d* = 0.4, and 80% power to detect significant differences in effort-based decision making between conditions as well as a linear regression model with a two-tailed alpha of *p* < .05, a moderate effect size of Cohen’s *f*^2^ = 0.16 and 80% power to detect significant brain-behavior associations. All participants were cognitively unimpaired at enrollment, aged 55 years or older, and had a first-degree family history of AD. Participants were enrolled in an optional sub-study, a randomized controlled trial aimed at increasing physical activity. The data analyzed in the present study were collected at baseline, prior to intervention onset. Details of the intervention and eligibility criteria have been reported previously.^68^

### Materials

#### The MacArthur Scale of Subjective Social Status

A scale used for assessing subjectively perceived socioeconomic status (SES).^69^ The scale presents a picture of a ladder with 10 rungs and participants are told to “imagine that the ladder represents where people stand in society.” The top rung (10) represents people who are the “best off” while the bottom rung (1) represents people who are the “worst off”. Participants are asked to select a number between 1 and 10 that best represents the rung where they think they stand in relation to others in society.

#### Patient Health Questionnaire-9

A questionnaire that consists of 9 items used for assessing symptoms of depression.^70^ Items ask how often over the past two weeks the respondent has been bothered by problems, such as “little interest or pleasure in doing things” and “feeling down, depressed, or hopeless”. Each item is scored from 0 to 3, with 0 = Not at all and 3 = Nearly every day. Scores range from 0 to 27, with higher scores indicating more severe symptoms of depression.

#### Repeatable Battery for the Assessment of Neuropsychological Status

Cognitive functioning was assessed using the Repeatable Battery for the Assessment of Neuropsychological Status,^71^ a standardized neuropsychological battery that evaluates five domains: Immediate Memory, Visuospatial/Constructional Ability, Language, Attention, and Delayed Memory. Domain and total index scores are age-normed standardized scores with a mean of 100 and a standard deviation of 15, with higher scores indicating better cognitive performance.

#### Effort-Based Decision-Making Task

Effort-based decision making was assessed using a task consisting of 120 trials in which participants decided whether to exert physical effort for monetary rewards.^72^ On each trial, participants were presented with a reward amount ($0.50–$3.00), reward type (prosocial or self-oriented), and required effort level. In the self-oriented condition, participants decided whether to exert effort to earn money for themselves, while in the prosocial condition, they decided whether to exert effort to donate money to a charity of their choosing. On each trial, participants were presented with a reward and the amount of effort required to obtain that reward, represented by the height of a red bar corresponding to one of five effort levels (3, 20, 37, 54, or 70 key presses). Participants indicated whether they accepted or rejected the offer by pressing ‘Y’ or ‘N’ on their keyboard. On one third of accepted trials, participants completed the required effort by alternating between the left and right arrow keys on their keyboard to raise the red bar to the height of a blue arrow. Participants were informed that one successfully completed trial would be randomly selected for payout.

**Figure 1.**
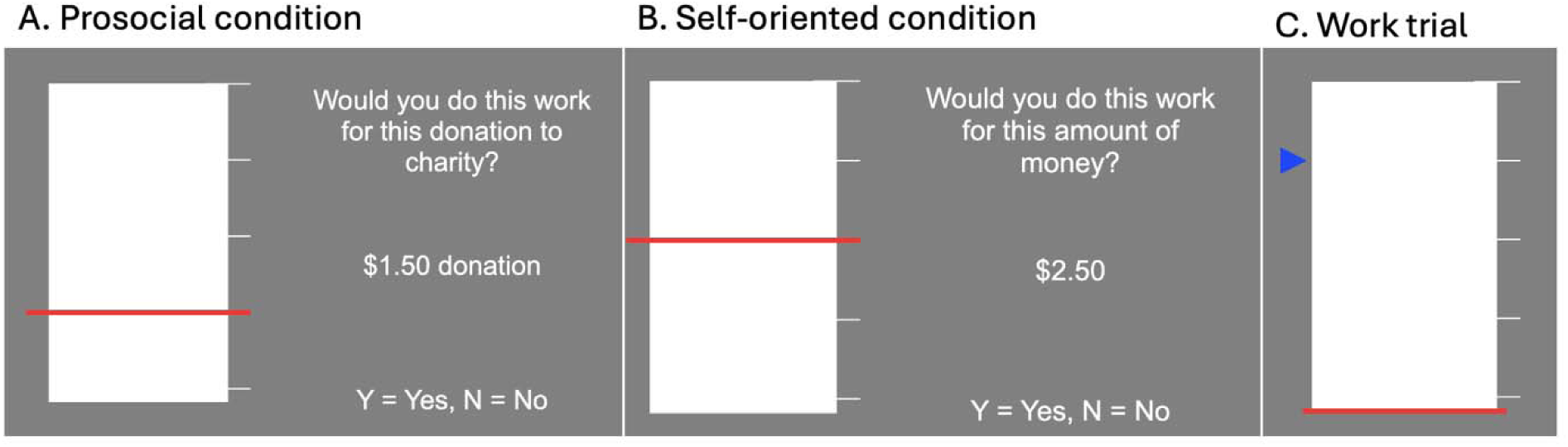
The effort-based decision-making task. The task consists of a prosocial (panel A) and self-oriented (panel B) reward condition. On each trial, participants accept or reject completing a certain amount of physical effort with a Y/N keyboard press. The amount of effort is represented by a red line at different heights, with a higher line requiring a greater amount of button presses. Prosocial or self-oriented monetary rewards vary between $0.50 - $3.00 on each trial. Thirty percent of trials are followed by work trials (Panel C) where participants must alternate between the left and right arrow keys to gradually move the red line up to the height of the blue arrow.

#### Procedure

Multi-echo resting-state functional magnetic resonance imaging (fMRI) data were collected between 2021 and 2022, and behavioral data were collected between 2021 and 2024. Participants were recruited from the PREVENT-AD cohort via phone, email, and mailed invitations. Eligible participants attended a videoconference session during which study procedures were explained and informed consent was obtained in accordance with the Declaration of Helsinki. Participants completed an online questionnaire battery administered via REDCap, followed by the effort-based decision making task.

#### APOE Genotyping

All participants were genotyped for APOE through a blood draw. DNA was isolated from 200 μl of the blood sample using the QIASymphony apparatus and the DNA Blood Mini QIA kit (Qiagen, Valencia, CA, USA). APOE gene variant was determined using pyrosequencing with PyroMark Q96 (Qiagen, Toronto, ON, Canada). *APOE4* carriership was defined as the presence of at least one ε4 allele.

#### FMRI Acquisition and Preprocessing

Participants were scanned on a 3T Siemens Magnetom Prisma MRI scanner with a 32-channel head coil (Siemens Medical Solutions, Erlangen, Germany). T1-weighted structural images were acquired using an MPRAGE sequence (1 mm isotropic voxels; TR = 2300 ms; TE = 2.96 ms; TI = 900 ms; flip angle = 9°; FOV = 256 mm; phase encode A-P; GRAPPA factor = 2; BW = 625 Hz/px). Resting-state fMRI data were acquired using a multi-echo echo-planar imaging (EPI) sequence during a single 10 min 24 s eyes-closed run (3 mm isotropic voxels; TR = 1000 ms; TE1 = 12 ms, TE2 = 30.11 ms, TE3 = 48.22 ms; flip angle = 50°; FOV = 240 mm; phase encode A-P; BW = 2500 Hz/px; GRAPPA ×2 acceleration; 48 slices).

Anatomical and functional MRI data were preprocessed using fMRIPrep v22.1.1.^73^ Susceptibility-induced distortions were estimated from a B0 fieldmap generated from paired EPI reference images and corrected using topup (FSL 6.0.5.1).^74^ Motion and distortion parameters were estimated from the first echo, and all echoes were resampled in native space. The first four volumes of each echo were removed to mitigate non–steady-state effects using fslroi. Multi-echo denoising was performed on minimally preprocessed scanner-space data using tedana’s TE-dependence analysis pipeline, which estimates voxelwise T2* to optimally combine echoes and separate BOLD from non-BOLD components following the Kundu decision tree.^75,76^ Subsequent analyses used the optimally combined denoised time series. Functional images were coregistered to T1 images and normalized to MNI152NLin2009cAsym space using antsRegistration (ANTs v2.3.3), then spatially smoothed with a 4-mm FWHM Gaussian kernel in CONN.^77^ Data quality was assessed using temporal signal-to-noise ratio, framewise displacement (>0.50 mm), and DVARS outliers (threshold = 20). Participants with >10% contaminated volumes were excluded, resulting in removal of three participants.

### Statistical Analysis

#### Behavioral

A generalized linear mixed-effects model was used to examine the influence of condition, reward, effort, and their interactions on the yes/no decision to exert effort using the *glmer* function from the lme4 package in R. Decision (yes or no) was coded as a binary outcome variable, and condition (prosocial vs. self-oriented), reward amount, effort level, and their interactions were included as fixed effects. Age, sex, years of education, SES, *APOE4* status, MCI status, and depression were included as covariates of non-interest in all models.

Robust linear mixed-effects models were conducted to examine the influence of condition, reward, effort, and decision (yes or no) and their interactions (fixed effects) on decision response times (outcome) as well as the influence of condition (fixed effect) on response vigor (outcome). The r*lmer* function from the robustlmm package in R was used for the robust linear mixed-effects models. Vigor was defined as the number of button presses per second on completed work trials. To examine differences between *APOE4* carriers and non-carriers in yes/no decisions, response times, and vigor, additional models were conducted with *APOE4* status included as an interaction term to the models. Subject-level random intercepts were included in all models. Post-hoc comparisons were performed using the emmeans package in R with a Bonferroni correction to adjust for multiple comparisons.

#### Computational Modelling

To characterize the maximum effort participants were willing to exert at each reward level, individual effort–reward curves were modeled using linear, sigmoid, and Weibull functions.^72^ Models were fit separately for each participant and condition using variational Bayes model inversion implemented in the VBA toolbox (MATLAB R2023b).^78^ Curve fitting was conducted within a mixed-effects framework to reduce the influence of outliers by iteratively estimating a Gaussian population-level prior from participant-level posterior estimates until convergence of group-level model evidence. Because both prosocial and self-oriented conditions contributed to the shared group prior, this procedure likely provided conservative estimates of condition differences by shrinking individual parameters toward the group mean.

The three functions used to map reward magnitude (x) onto maximum effort exerted (y) were:

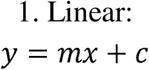

where *m* is the slope and *c* is the intercept.

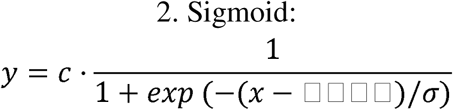

where *bias* determines the horizontal shift of the curve (tendency to avoid effort at a given reward) and σ reflects reward sensitivity (change in effort per unit reward).

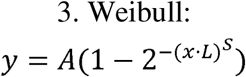

where *L* represents the minimum reward needed to initiate effort (latency), *S* controls how abruptly effort increases with reward (reward sensitivity), and *A* determines the maximum achievable effort.

Random-effects Bayesian model comparison was used to determine the best-fitting model based on protected exceedance probability. To confirm model equivalence across conditions, between-condition random-effects model comparison was conducted on the same model set. Parameter reliability was assessed using a simulation–recovery procedure with 1,000 parameter sets sampled from the group-level posterior distribution. Synthetic datasets were generated using these parameters and re-inverted using the same procedure, and recovered parameters were correlated with the original sampled values using product-moment correlations. Parameter estimates from the winning model were then extracted for each participant and condition. Linear mixed-effects models (lmer, lme4 package in R) tested condition effects on parameter estimates with subject-level random intercepts, and additional models examined condition × APOE4 interactions.

#### Resting-state fMRI

Resting-state fMRI analyses were conducted using the CONN toolbox (RRID:SCR_009550; release 22.v2407).^77^ For model parameters showing significant condition differences, within-subject difference scores (e.g., bias_self-oriented_ – bias_prosocial_) were computed to index greater prosocial relative to self-oriented effort-based decision making. Seed-to-voxel analyses examined associations between these difference scores and rsFC. Seeds included the vmPFC and ACC, selected *a priori* based on prior evidence implicating these regions in prosocial behavior. The vmPFC seed was defined using a meta-analytic mask of subjective value processing and the ACC seed using an anatomical mask previously associated with prosocial effort-based decision making.^26,39^ Functional connectivity was estimated as Fisher-transformed correlations between seed and voxel BOLD time series using CONN’s weighted general linear model. Cluster-level inference used a voxel threshold of p < .001 and cluster-level FDR correction of p < .05 based on Gaussian random field theory.

ROI-to-ROI analyses examined rsFC differences between APOE4 carriers and non-carriers within a predefined reward circuit comprising the VTA, NAcc, dACC, and vmPFC. Regions were defined using established atlases (VTA probabilistic atlas, Harvard–Oxford for bilateral NAcc, Desikan–Killiany for dACC, and meta-analytic vmPFC mask).^26,79–81^ Functional connectivity was estimated using Fisher-transformed correlations within CONN’s weighted-GLM framework. Group-level GLMs examined *APOE4* carrier status as the effect of interest while controlling for the same covariates as seed-to-voxel analyses, with FDR correction applied across connections (p < .05). Significant connections were followed up with exploratory linear and robust regression analyses (lm and rlm in R) examining associations with effort-based decision-making outcomes.

Functional connectivity multivariate pattern analysis (fc-MVPA) was used to identify data-driven associations between effort-based decision making parameter difference scores and rsFC, as well as rsFC differences between *APOE4* carriers and non-carriers. Fc-MVPA estimated the first five eigenpatterns capturing principal axes of inter-individual variability in whole-brain functional connectivity. Eigenpatterns and subject-specific eigenpattern scores were derived using singular value decomposition of seed-to-voxel connectivity matrices. Group-level voxelwise GLMs were estimated on eigenpattern scores using the same inference framework and correction procedures as the seed-to-voxel analyses. Clusters exceeding k > 50 voxels were followed up with seed-to-voxel analyses using identical models as described above.

To assess robustness to potential outliers, significant effects were followed up using post-hoc robust regression analyses with M-estimation implemented in the *rlm* function from the MASS package in R (see Supplementary Materials). 95% confidence intervals for regression coefficients were obtained using nonparametric bootstrapping with 1,000 resamples using R’s boot package.

## Results

### Participant Demographics

129 participants were screened for eligibility, with 61 excluded as a result of not meeting inclusion criteria and 16 deemed eligible who declined participating, resulting in a final sample of *N* = 52 included in final behavioral analyses. Three additional participants were excluded from neuroimaging analyses due to motion-related artifacts. Demographic, psychiatric, and cognitive information for the full sample, and for each *APOE* group, is shown in Table 1. There were no statistically significant differences between the *APOE* groups in any characteristics.

**Table 1.** Demographic characteristics of the full sample (*M* ± *SD*).

| | Total Sample<br>($N = 52$) | APOE4 carriers<br>($n = 18$) | APOE4 non-carriers<br>($n = 34$) |
| --- | --- | --- | --- |
| Age | 68.48 ± 4.35 | 67.5 ± 5.48 | 69 ± 3.59 |
| Sex | Female = 38<br>Male = 14 | Female = 14<br>Male = 4 | Female = 24<br>Male = 10 |
| Socioeconomic status (MacArthur Scale of Perceived Social Status) | 6.71 ± 1.49 | 6.72 ± 1.49 | 6.7 ± 1.51 |
| Education (years) | 16.04 ± 3.17 | 15.5 ± 4.18 | 16.32 ± 2.51 |
| MCI status | CU = 41<br>MCI = 11 | CU = 15<br>MCI = 3 | CU = 26<br>MCI = 8 |
| Depression (PHQ-9) | 2.38 ± 2.53 | 2.56 ± 2.25 | 2.29 ± 2.69 |
| RBANS Total | 102.13 ± 8.34 | 102.5 ± 9.63 | 101.94 ± 7.72 |
| RBANS Immediate memory | 104.9 ± 10.13 | 108 ± 9.26 | 103.26 ± 10.31 |
| RBANS Visuospatial/Constructional | 85.58 ± 9.71 | 84.5 ± 12.06 | 86.15 ± 8.35 |
| RBANS Language | 98.29 ± 9.81 | 98.83 ± 12.21 | 98 ± 8.46 |
| RBANS Attention | 111.15 ± 15.56 | 110.89 ± 18.73 | 111.29 ± 13.9 |
| RBANS Delayed memory | 101.73 ± 7.75 | 102.89 ± 7.68 | 101.12 ± 7.83 |

*Note.* CU = cognitively unimpaired; MCI = mild cognitive impairment; RBANS = Repeatable Battery for the Assessment of Neuropsychological Status.

### Behavioral

Results of the generalized linear mixed-effects model showed that participants accepted more prosocial than self-oriented reward trials (β = 0.682, *SE* = 0.313, *z* = 2.18, *p* = .029, *OR* = 1.98, 95% *CI* [1.07, 3.65]). A significant reward value × effort level interaction was observed across conditions (β = 0.01, *SE* = 0.002, *z* = 2.07, *p* = .039, *OR* = 1.01, 95% *CI* [1.00, 1.01]).

Post-hoc contrasts showed a decrease in effort acceptance for small increases in effort when rewards were low, while at higher reward values, only the highest effort level (70 presses) significantly reduced acceptance relative to lower effort levels (3 or 20 presses). Main effects indicated that higher effort significantly reduced effort acceptance (β = −0.038, *SE* = 0.01, *z* = −7.58, *p* < .001, *OR* = 0.96, 95% *CI* [0.95, 0.97]) and higher reward significantly increased effort acceptance (β = 1.10, *SE* = 0.123, *z* = 8.94, *p* < .001, *OR* = 3.01, 95% *CI* [2.37, 3.84]).

Robust linear mixed-effects models revealed significantly longer decision response times in the prosocial compared to the self-oriented condition (β = -0.54, *SE* = 0.21, *z =* -2.54, *p* = .011) and as the level of effort required decreased (β = -0.02, *SE* = 0.01, *z* = -2.30, *p* = .021). There was no effect of condition on response vigor (*p* > .05).

**Figure 2.**
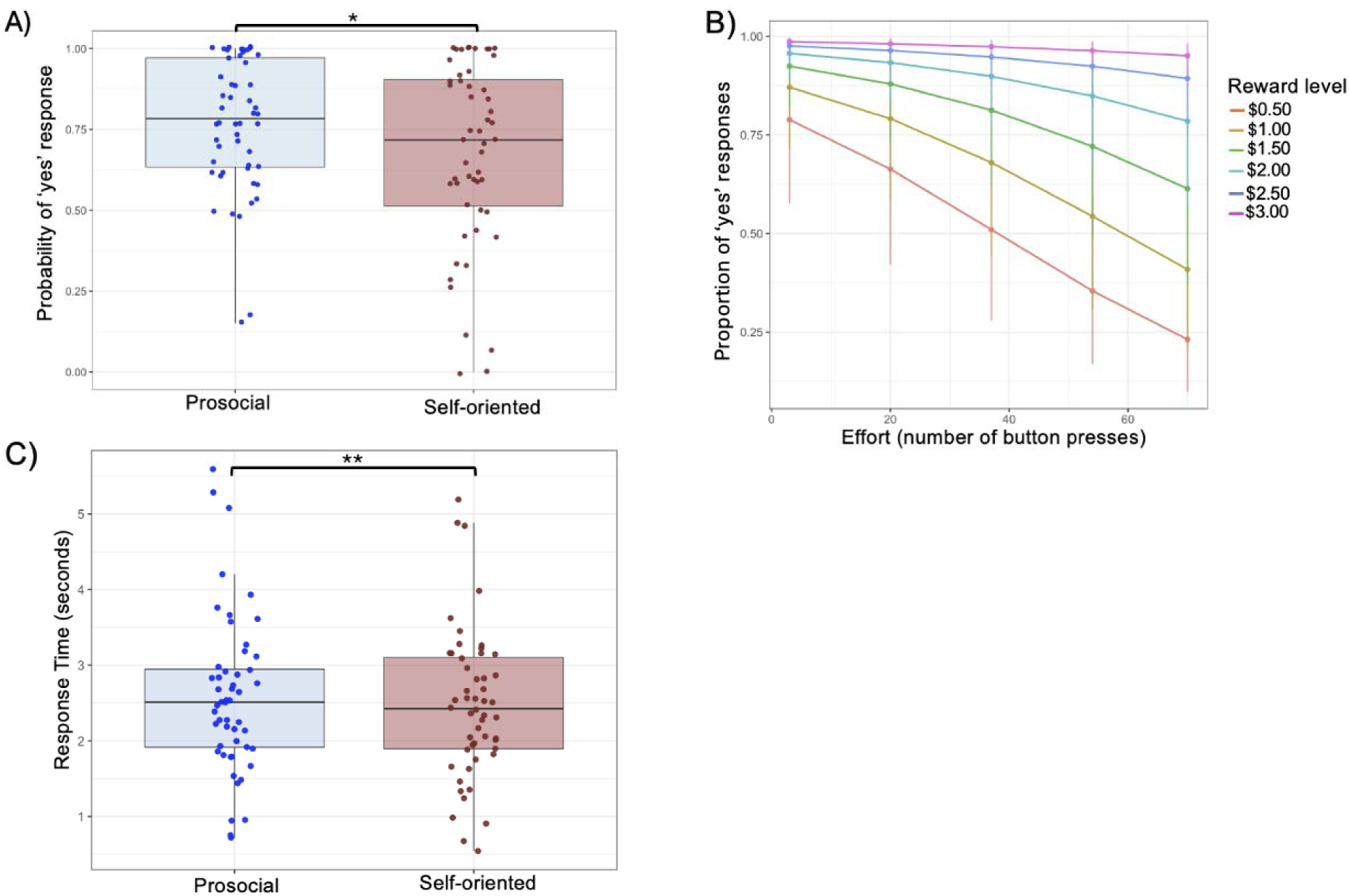
Effort acceptance and decision response times by condition. The probability of accepting effort is significantly higher for prosocial compared to self-oriented rewards (Panel A) and effort acceptance is maintained with increasing effort with higher reward values (Panel B). Decision response times were significantly higher in the prosocial compared to self-oriented reward condition (Panel C), \**p* < .05, \*\**p* < .01.

### Computational Modelling

Random-effects Bayesian model comparison indicated that the sigmoid function provided the best fit to reward-effort discounting curves (protected exceedance probability > 0.999 and estimated model frequency = 0.976). Bayesian model selection further showed no difference in model fit between the prosocial and self-oriented reward conditions (protected exceedance probability = 1.00 for model equivalence). The sigmoid model provided a good account of individual effort-reward functions across participants (mean *R*^2^ = 0.883, *SD* = 0.185 for prosocial condition; mean *R*^2^ = 0.896, *SD* = 0.153 for self-oriented condition). The correlations between simulated and recovered parameter estimates for *N* = 1000 randomly drawn samples were *r* = .627 for sigma and *r* = .692 for bias, indicating acceptable reliability.

Results of the linear mixed-effects models on sigmoid model parameters showed that bias away from effort was significantly higher in the self-oriented compared to prosocial condition (β = 5.98, *SE* = 2.13, *t* = 2.81, *p* = .007). There was no significant effect of condition on sigma/reward insensitivity (*p* > .05).

**Figure 3.**
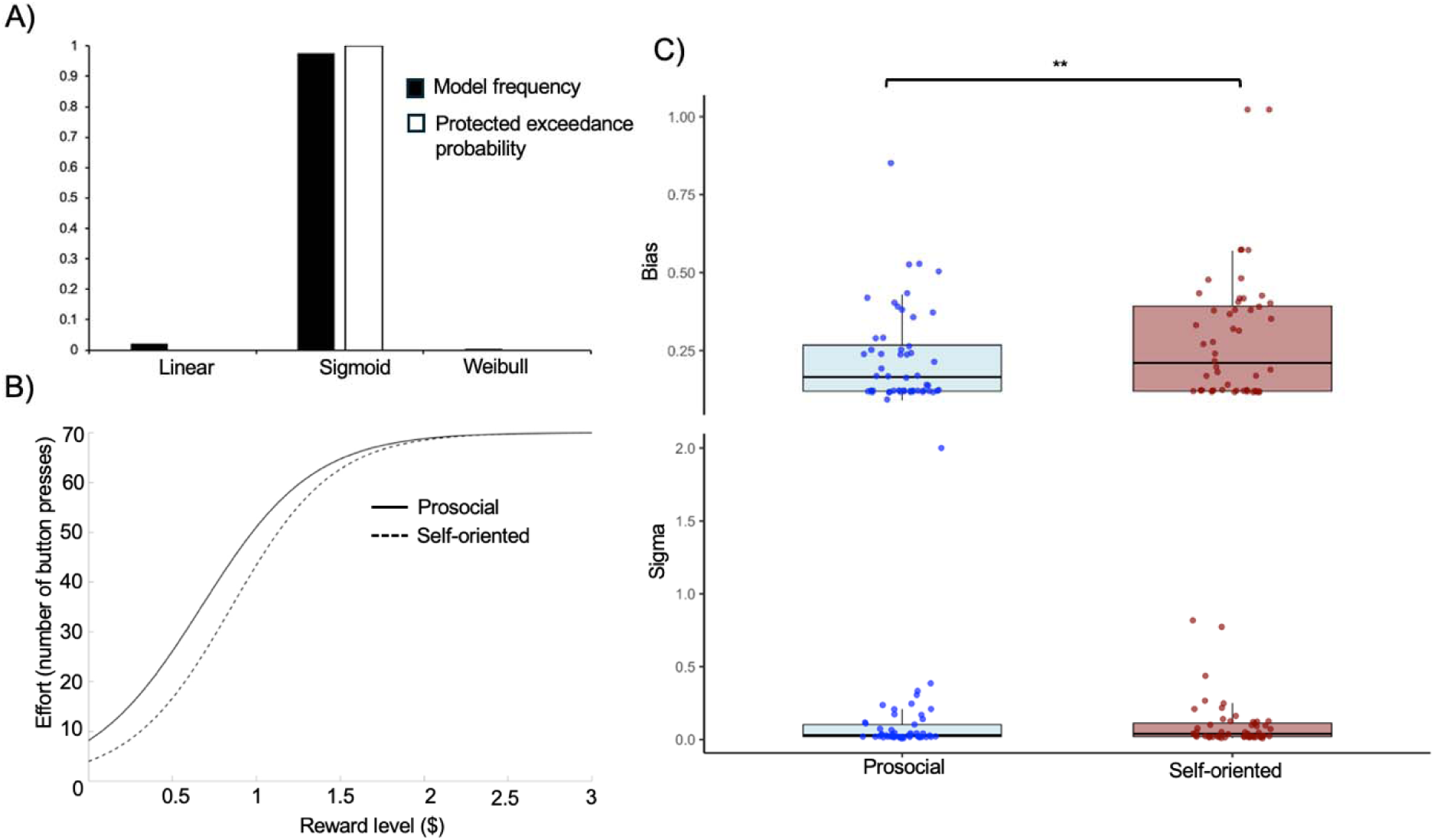
Results of modelling the reward-effort relationship. A sigmoid function represented a better overall fit to the reward-effort data than a linear or Weibull model (Panel A). Bias away from effort (right translation of the curve) was significantly higher in the self-oriented compared to prosocial reward condition while there were no differences between conditions in reward insensitivity/sigma (slope of the curve) (Panels B and C). \*\**p* < .01

### Behavioral Differences between *APOE4* Carriers and Non-Carriers

A significant condition × *APOE4* status interaction was observed for response vigor (β = -0.62, *SE* = 0.18, *z* = -3.46, *p* < .001). Post-hoc comparisons (Table S4) indicated higher vigor for prosocial relative to self-oriented rewards in *APOE4* carriers, with no differences between conditions among non-carriers. *APOE4* carriers also exhibited greater vigor than non-carriers for prosocial, but not self-oriented, rewards. In addition, *APOE4* status was associated with greater bias away from effort expenditure across conditions, with carriers showing higher bias than non-carriers (β = 0.10, *SE* = 0.01, *t* = 2.15, *p* = .037).

**Figure 4.**
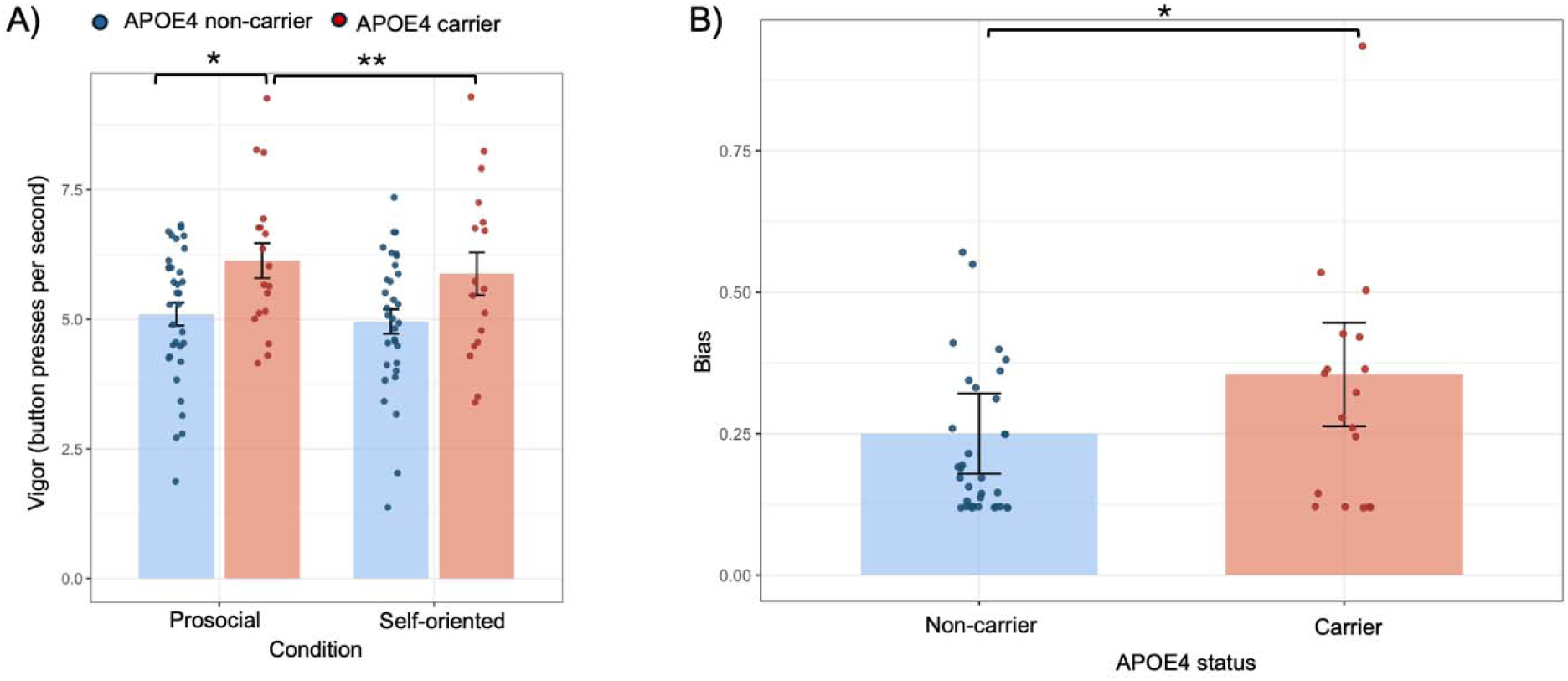
Differences in effort-based decision making by *APOE4* carriership. Vigor was significantly higher for *APOE4* carriers than non-carriers when rewards were prosocial (Panel A) and bias away from effort expenditure was higher for *APOE4* carriers compared to non-carriers across both conditions (Panel B). \*\**p* < .01, \**p* < .05

### Resting-State fMRI Results

#### Region of Interest Analyses

Bias difference scores (self-oriented − prosocial) were associated with enhanced rsFC between the vmPFC seed and the left dorsomedial PFC (*t*(39) = 5.22, peak MNI = [0, 38, 44], k = 86). Bias difference scores were also associated with enhanced rsFC between the ACC seed and two clusters in the left inferior frontal gyrus, pars triangularis (cluster 1: *t*(39) = 6.32, peak MNI = [−42, 26, 14], k = 149; cluster 2: *t*(39) = 4.66, peak MNI = [−54, 26, 20], k = 47). In contrast, bias difference scores were associated with reduced rsFC between the ACC seed and the right superior lateral occipital cortex (*t*(39) = −6.07, peak MNI = [30, −78, 44], k = 108), right superior parietal lobule (*t*(39) = −4.69, peak MNI = [30, −52, 60], k = 51), right anterior middle/inferior temporal gyrus (*t*(39) = −6.43, peak MNI = [60, −8, 8], k = 50), and right dorsolateral PFC (*t*(39) = −6.04, peak MNI = [24, 12, 42], k = 48).

**Figure 5.**
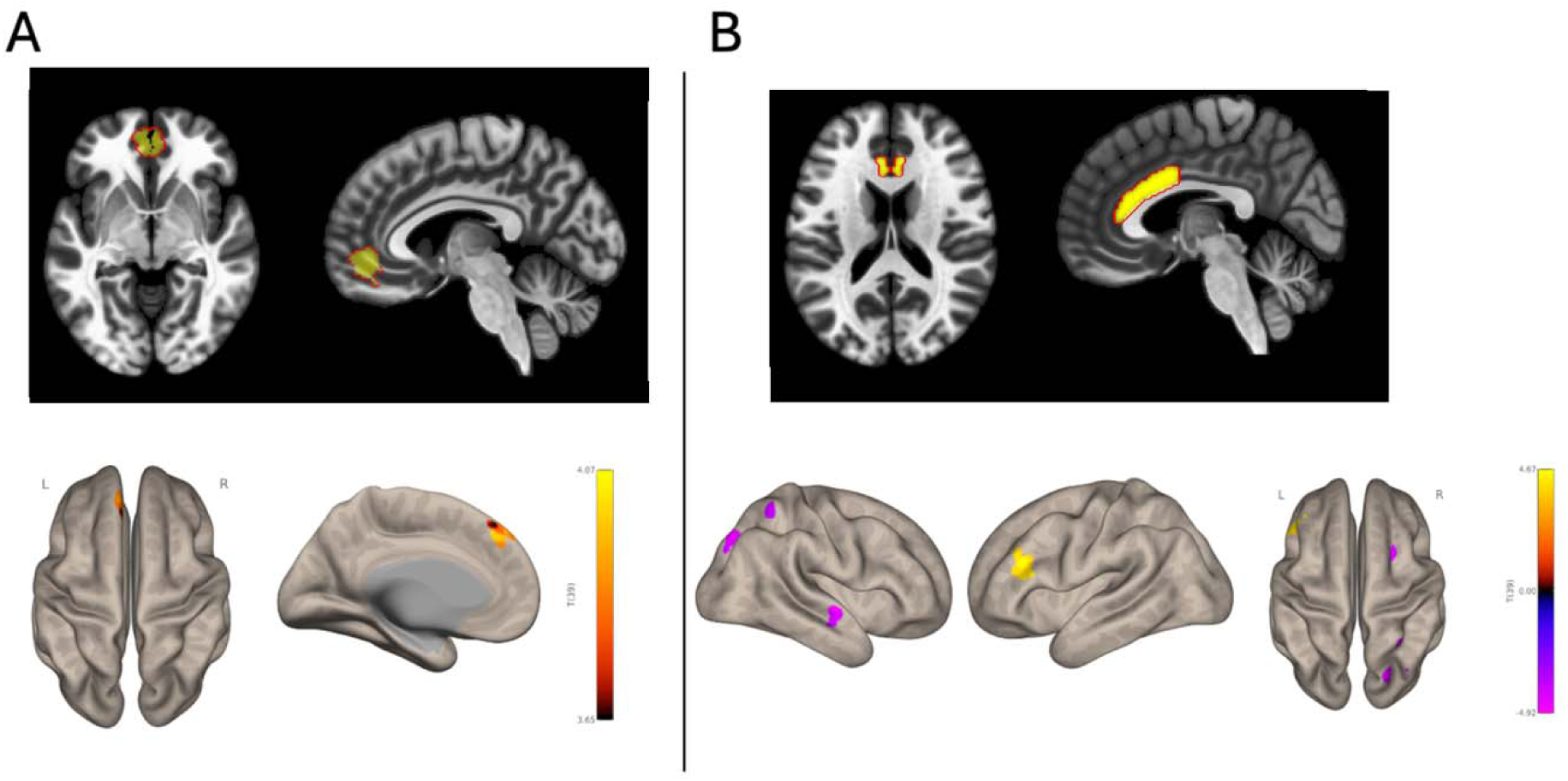
Seed-to-voxel resting-state functional connectivity associated with bias towards prosocial effort. Higher bias towards effort expenditure for prosocial relative to self-oriented rewards was associated with enhanced rsFC between the ventromedial prefrontal cortex and left dorsomedial prefrontal cortex (panel A) and the gyrus of the anterior cingulate cortex (panel B) with the left inferior frontal gyrus (positive rsFC) and right superior lateral occipital cortex, right superior parietal lobule, right anterior middle/inferior temporal gyrus, and right dorsolateral prefrontal cortex (anticorrelated rsFC). Results are overlaid on a standard MNI template and corrected for multiple comparisons.

The ROI-to-ROI analyses revealed that *APOE4* carriers, relative to non-carriers, had reduced rsFC between the right NAcc and dACC (*t*(40) = -3.55, *p*-FDR = .01). Moreover, lower right NAcc-dACC rsFC was positively associated with bias away from effort expenditure across both conditions, β = -0.257, *SE* = 0.11, *t*(40) = -2.32, *p* = .025.

#### Multivariate Pattern Analyses

Fc-MVPA revealed that bias difference scores (self-oriented - prosocial) were associated with three vmPFC clusters across all participants: a posterior vmPFC/subcallosal cluster spanning both hemispheres (labelled 1 in Fig. 6A; *F*(5,35) = 9.09, peak MNI = [0, 4, 28], k = 676), an anterior vmPFC cluster extending into the bilateral medial frontal poles (labelled 3 in Fig. 7A; *F*(5,35) = 5.05, peak MNI = [2, 56, −28], k = 298), and a right posterior vmPFC cluster (labelled 2 in Fig. 6A; *F*(5,35) = 8.05, peak MNI = [18, 50, 10], k = 96).

**Figure 6.**
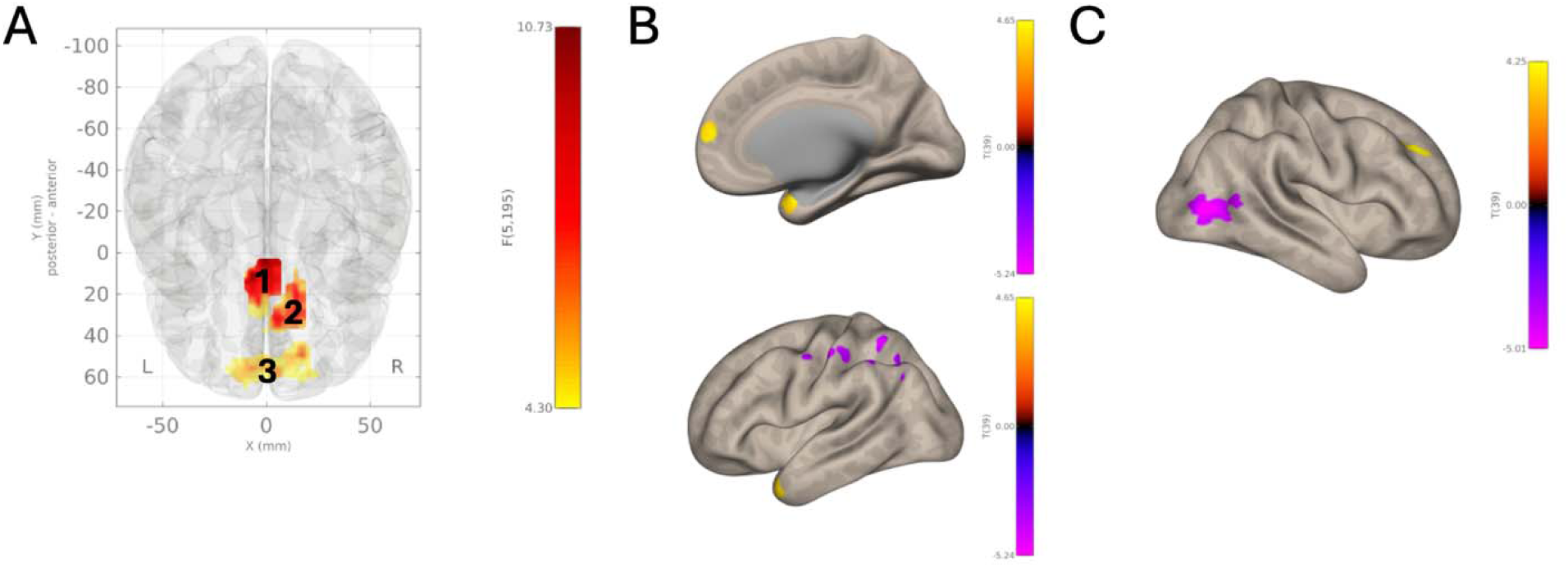
Whole-brain functional connectivity multivariate pattern analyses associated with bias towards prosocial effort. Panel A shows three seeds located in the ventromedial prefrontal cortex. Panel B shows resting-state functional connectivity between vmPFC seed 3 with the bilateral temporal poles and right frontal pole (positive connectivity) and with the left lateral occipital cortex / superior parietal lobule, left postcentral gyrus, and left temporoparietal junction (negative connectivity). Panel C shows resting-state functional connectivity between vmPFC seed 1 with the right dorsolateral prefrontal cortex (positive connectivity) and right lateral occipital cortex (negative connectivity). No significant effects were found with seed 2.

Post-hoc seed-to-voxel analyses using these clusters as seeds showed that bias difference scores were associated with enhanced rsFC between the posterior vmPFC/subcallosal seed (seed 1 in Fig. 7A) and the right dorsolateral PFC (*t*(39) = 5.78, peak MNI = [28, 44, 36], k = 61), and reduced rsFC with the right lateral occipital cortex (*t*(39) = −7.02, peak MNI = [52, −72, 8], k = 268). Bias difference scores were also associated with enhanced rsFC between the anterior vmPFC/frontal pole seed (seed 3 in Fig. 7A) and the right temporal pole (*t*(39) = 6.64, peak MNI = [32, 4, 26], k = 199), left temporal pole (*t*(39) = 5.02, peak MNI = [−46, 20, −34], k = 95), and right frontal pole (*t*(39) = 4.96, peak MNI = [12, 56, 14], k = 55), as well as reduced rsFC with the left lateral occipital cortex/superior parietal lobule (*t*(39) = −6.14, peak MNI = [−36, −52, 54], k = 93), left postcentral gyrus (*t*(39) = −6.61, peak MNI = [−50, −16, 50], k = 65), and left temporoparietal junction (*t*(39) = −5.64, peak MNI = [−54, −40, 54], k = 54). Fc-MVPA did not identify any significant clusters differentiating *APOE4* carriers from non-carriers.

## Discussion

This study examined effort-based decision making for prosocial versus self-oriented rewards and the rsFC underlying prosocial effort-based decision making in older adults at risk for AD. Given evidence for heightened social motivation in aging and relatively preserved socioemotional functioning in early AD, we tested whether prosocial incentives enhance effort-based decision making and whether these processes are altered in carriers of the *APOE4* allele, a genetic risk factor for AD.^9,12,22,50^ Participants showed greater acceptance of effort for prosocial than self-oriented rewards and took longer to reach a decision threshold for prosocial rewards.

Greater bias toward prosocial effort was associated with enhanced vmPFC–dmPFC and ACC–left inferior frontal gyrus rsFC, and reduced ACC connectivity with lateral occipital cortex, superior parietal lobule, inferior/middle temporal gyrus, and dlPFC. Compared with non-carriers, APOE4 carriers showed reduced dACC–NAcc connectivity, which was associated with greater bias away from effort across conditions.

Older adults showed greater acceptance and bias towards prosocial compared to self-oriented effort, consistent with evidence that aging is associated with heightened prosocial behavior and social motivation.^9,12,19^ These findings partially align with prior work showing reduced effort discounting for prosocial rewards in older relative to younger adults, though previous within-group comparisons in older adults reported lower acceptance of effort for prosocial compared to self-oriented rewards.^9^ Such discrepancies may reflect methodological differences, including the type and frequency of effort and the subjective value of the reward. In the present study, effort involved keyboard presses completed on one third of accepted trials, whereas prior work required squeezing a dynamometer on all accepted trials, potentially making effort in the current task less costly or more uncertain. Additionally, rewards were charitable donations, which may be more meaningful to older adults than abstract points or credits. This interpretation is consistent with evidence that prosocial behavior in aging increases when effort costs are lower and outcomes align with emotionally meaningful goals.^82^

Consistent with prior work, vigor across the whole sample did not differ between prosocial and self-oriented conditions.^9^ This suggests that at-risk older adults exert as much effort to obtain a prosocial reward compared to a reward for themselves and that prosocial incentives primarily enhance the decision to exert effort rather than the magnitude of effort exerted once a decision has been made. Similarly, the slope of the effort–reward sigmoid function did not differ between prosocial and self-oriented conditions. This suggests that older adults’ decisions were equally sensitive to increases in reward size regardless of who benefited. However, the monetary amounts used in this task were small ($0.50–$3.00), which may have limited the extent to which reward-size differences between conditions influenced acceptance of effort expenditure. Future studies should therefore examine these processes in more ecologically valid contexts to determine whether the stability of reward sensitivity observed here generalizes to real-world decisions.

Contrary to our expectations, older adults took longer to reach a decision threshold for prosocial compared to self-oriented rewards. Longer response times may reflect increased deliberative processing during decision making.^83–85^ Thus, one possibility is that prosocial effort is perceived as more cognitively demanding because the benefits are less immediate and self-directed, requiring greater integration of value, effort, and social cognitive processing. Prosocial decisions may therefore recruit additional deliberative processes such as inhibitory control, value integration, and mentalizing.^83,86^ Future work should directly test this interpretation by manipulating cognitive load, temporal delay, or mentalizing demands during prosocial versus self-oriented effort-based decision-making to determine how these processes influence effort-based decisions.

Beyond condition differences, effort acceptance generally declined as effort demands increased and reward magnitude decreased, consistent with effort discounting. However, at higher levels of reward magnitude, effort acceptance remained increasingly more stable across effort levels, suggesting that sufficiently large incentives can help override increases in effort costs for both reward types. These findings have important practical implications, indicating that effort sensitivity in older adults at risk for AD is modifiable. Small rewards may be insufficient for motivating effort when tasks become too demanding and increases in effort without commensurate increases in reward may contribute to behavioral disengagement. Strategically deploying larger or more salient incentives during high-effort periods, such as at the initiation of a behavior change program, may provide a means to help sustain engagement.

At the neural level, both seed-to-voxel and fc-MVPA analyses converged on the vmPFC, a node of the reward and default mode networks, as a key hub underlying individual differences in bias towards prosocial relative to self-oriented effort. Meta-analytic work suggests a rostrocaudal organization of vmPFC, with posterior regions integrating affective and motivational relevance and anterior regions supporting valuation, social cognition, and action selection.^87^ Bias towards prosocial effort was associated with stronger rsFC between the vmPFC seed implicated in valuation^26^ and dmPFC, a core node of the default mode network involved in perspective taking, mentalizing, and self-referential processing.^88,89^ Prior work demonstrates vmPFC–dmPFC co-activation during social decision making, with dmPFC activity scaling with cognitive and physical effort devoted to valued goals.^88,90^ Enhanced vmPFC–dmPFC coupling may therefore reflect more effective integration of valuation and social-cognitive representations, a hypothesis to be tested in future work.

Higher bias toward prosocial effort was associated with increased rsFC between vmPFC clusters identified by fc-MVPA and the right dlPFC and bilateral temporal poles. The dlPFC, a hub of the frontoparietal control network involved in planning and goal-directed behavior,^91,92^ has been linked to goal simulation, generativity, and prioritizing options that benefit others over the self when co-active with the vmPFC.^25,93,94^ Moreover, the vmPFC and temporal poles form part of a limbic subsystem of the default mode network supporting empathy, moral reasoning, and valuation of others’ outcomes.^95–97^ In contrast, greater bias towards prosocial effort was associated with anticorrelated rsFC between vmPFC clusters and regions of perceptual and attention networks, including the lateral occipital cortex, superior parietal lobule, postcentral gyrus, and left temporoparietal junction.^87,98–101^ Anticorrelation between default mode and perceptual–attentional networks is often reduced in aging and AD and its preservation is linked to better cognition.^102,103^ This pattern may reflect a shift away from externally driven processing toward internally guided valuation and socioemotional representations during prosocial decision making. This interpretation aligns with evidence that aging is associated with a shift from exploratory to exploitative decision strategies that prioritize emotionally meaningful goals.^104^

Bias towards prosocial compared to self-oriented effort was also associated with rsFC to the ACC, a region central to monitoring others’ outcomes, encoding social value, and prosocial effort-based decision-making.^35,38^ Stronger rsFC between ACC and left IFG and reduced rsFC between the ACC with regions of the frontoparietal and attentional-perceptual networks (e.g., lateral occipital cortex, superior parietal lobule, inferior/middle temporal gyrus, dlPFC) was associated with higher bias towards prosocial effort. Parts of the left IFG have been hypothesized to include mirror neurons crucial for the social perception of emotions.^105,106^ The IFG has been shown to co-activate with the ACC during semantic processing, emotion regulation, and cognitive control,^107,108^ processes crucial for social behavior. Reduced coupling between ACC and regions of attentional-perceptual/frontoparietal networks may indicate that prosocial effort decisions rely less on deliberative control and external action planning than self-oriented decisions, aligning with evidence that ACC–frontoparietal connectivity increases during cognitively demanding tasks but decreases at rest when motivational priorities guide action selection.^109^

Comparisons between *APOE4* carriers and non-carriers revealed that carriers showed a greater overall bias away from exerting effort across reward types yet exhibited heightened vigor once engaging in effort for prosocial relative to self-oriented rewards. This suggests *APOE4* carriers accept smaller amounts of effort for a given reward value than non-carriers but display enhanced energization to obtain prosocial rewards after they have been accepted. One possible explanation that warrants future investigation is altered dopaminergic function in *APOE4* carriers, such as diminished tonic signalling supporting baseline motivation alongside relatively intact or enhanced phasic responses to salient, prosocial rewards that invigorate action.^110–113^ Consistent with this account, one animal study reported lower dopamine levels in the frontal cortex of *APOE4* mice compared to *APOE2* and *APOE3* mice.^114^ In addition, some studies suggest that vigor is supported by dopaminergic signalling in the nigrostriatal pathway comprising the substantia nigra and dorsal striatum.^115,116^ Thus, future studies should examine how *APOE4* carriership might affect rsFC of this circuit and its relationship with vigor.

*APOE4* carriers showed reduced rsFC between the dACC and right NAcc, with reduced rsFC associated with greater bias away from effort expenditure. Converging evidence from animal and human studies indicate that the dACC-NAcc circuit plays a critical role in integrating effort costs and reward valuation during motivated behavior, supporting choices toward high effort options and reducing reward thresholds required to engage in effort.^91,117^ Thus, disruptions in this circuit in *APOE4* carriers may impact how reward and effort are integrated to drive goal-directed behavior. Moreover, the present results were observed despite no group differences in global cognition and after controlling for MCI status, suggesting that subtle alterations in effort-based decision-making and reward circuits may precede overt cognitive impairment in *APOE4* carriers at a heightened risk for AD.

Despite the insights gained from this study, several limitations warrant consideration. First, the sample was primarily composed of female, Caucasian, highly educated older adults from Quebec, Canada with a family history of AD who were recruited from a longitudinal AD prevention cohort. As such, participants may have been especially research-engaged and socially motivated, which may limit generalizability to broader and more diverse older adult populations. Second, the within-subjects design may have increased social desirability or subject-expectancy effects. Because only older adults were included, it also remains unclear how these findings compare with younger adults, and direct age-group comparisons are needed to clarify lifespan changes in prosocial effort-based decision-making. Third, rewards were monetary, whereas many real-world prosocial behaviors are intrinsically rather than financially rewarding. Future work should therefore examine whether similar behavioral and neural patterns emerge for non-monetary prosocial acts. Finally, future studies should examine neuropathological mechanisms underlying APOE4-related differences in effort-based decision making, including potential contributions of inflammatory, amyloid, and neurotrophic markers.^118–120^

In summary, this study provides evidence that older adults at risk for AD accept more decisions to exert effort for prosocial than self-oriented rewards, with this preference associated with rsFC involving the vmPFC and ACC. Reduced rsFC within a reward circuit (dACC–NAcc) among APOE4 carriers suggests that this genetic risk may contribute to subtle alterations in motivation prior to cognitive decline. From a translational perspective, prosocial incentives may help maintain or enhance willingness to expend effort in at-risk older adults. Although APOE4 carriers accepted lower levels of effort overall, they showed greater vigor for prosocial relative to self-oriented rewards, suggesting that prosocial incentives may be particularly effective at energizing action in this population. Because prosocial behaviors such as volunteering, caregiving, and charitable giving are associated with improved well-being, social connectedness, and reduced dementia risk,^12,121,122^ leveraging prosocial rewards may represent a promising strategy to support goal-directed behavior and brain health in aging.

## Data availability

All data and materials can be made available upon request.

## Acknowledgements

We thank all the participants of the PREVENT-AD cohort for their time and effort. We also thank all students and research assistants who assisted with data collection. We would like to thank Claire Webster and Dr. Jessica Cooper for their valuable input on the project. We would also like to thank Margie Lachman for her leadership of the Boston Roybal Centre and Annie Le Bire and Joanna Masterlick for administrative support. We thank all the collaborators, consultants, and staff members from the PREVENT-AD Research Group. A complete list of the PREVENT-AD Research Group can be found at: https://preventad.loris.ca/acknowledgements/acknowledgements.php?date=.

## Funding

This research was undertaken thanks in part to funding from a National Sciences and Engineering Research Council of Canada (NSERC) Discovery Grant (DGECR-2022-00299), an NSERC Early Career Researcher Supplement (RGPIN-2022-04496), a Fonds de Recherche Santé Québec (FRQS) Salary Award, the Canada Brain Research Fund (CBRF), an innovative arrangement between the Government of Canada (through Health Canada) and Brain Canada Foundation, an Alzheimer Society Research Program (ASRP) New Investigator Grant, the Canadian Institutes of Health Research (CIHR), the Canada First Research Excellence Fund, awarded through the Healthy Brains, Healthy Lives initiative at McGill University, and the National Institutes of Health (P30 AG048785) to MRG. This research was also undertaken thanks in part to funding from the CIHR Doctoral Research Award: Canada Graduate Scholarship (199381) to CSW.

## Competing interests

The authors declare that they have no known competing financial interests or personal relationships that could have appeared to influence the work reported in this paper.

## Notes

### Competing Interest Statement

The authors have declared no competing interest.

### Funding Statement

This research was supported in part by a Natural Sciences and Engineering Research Council of Canada (NSERC) Discovery Grant (DGECR-2022-00299), an NSERC Early Career Researcher Supplement (RGPIN-2022-04496), a Fonds de Recherche Sante Quebec (FRQS) Salary Award, the Canada Brain Research Fund (CBRF), a partnership between the Government of Canada (through Health Canada) and the Brain Canada Foundation, an Alzheimer Society Research Program (ASRP) New Investigator Grant, the Canadian Institutes of Health Research (CIHR), the Canada First Research Excellence Fund awarded through the Healthy Brains, Healthy Lives initiative at McGill University, and the National Institutes of Health (P30 AG048785) to MRG. CSW was supported by a CIHR Doctoral Research Award: Canada Graduate Scholarship (199381).

### Author Declarations

IRB of McGill University gave ethical approval for this work.

### Summary of Updates

This version incorporates additional supplementary materials not present in the manuscript

