## Supplementary Materials for "The neural basis of prosocial effort-based decision making in older adults at risk for Alzheimer’s disease"

**Supplementary Material**

**Behavioral Results**

**Table** **S1** Post-hoc comparisons between reward levels by effort magnitude on choice data.

| Comparison | Odds Ratio | 95% CI | p (Bonferroni corrected) |
| --- | --- | --- | --- |
| $0.50 vs. $1, Effort = 3 | 3.79 | 1.66, 8.63 | <.001 |
| $0.50 vs. $1.50, Effort = 3 | 2.36 | 1.08, 5.13 | .018 |
| $0.50 vs. $2.00, Effort = 3 | 5.68 | 2.36, 13.68 | <.001 |
| $0.50 vs. $2.50, Effort = 3 | 10.22 | 3.76, 27.76 | <.001 |
| $0.50 vs. $3.00, Effort = 3 | 11.45 | 4.15, 31.57 | <.001 |
| $1.00 vs. $1.50, Effort = 3 | 0.62 | 0.27, 1.45 | .999 |
| $1.00 vs. $2.00, Effort = 3 | 1.50 | 0.59, 3.83 | .999 |
| $1.00 vs. $2.50, Effort = 3 | 2.70 | 0.94, 7.71 | .083 |
| $1.00 vs. $3.00,  Effort = 3 | 3.02 | 1.04, 8.76 | .035 |
| $1.50 vs. $2.00,  Effort = 3 | 2.41 | 0.98, 5.92 | .061 |
| $1.50 vs. $2.50,  Effort = 3 | 4.34 | 1.57, 11.98 | <.001 |
| $1.50 vs. $3.00,  Effort = 3 | 4.86 | 1.73, 13.62 | <.001 |
| $2.00 vs. $2.50, Effort = 3 | 1.80 | 0.60, 5.37 | .999 |
| $2.00 vs. $3.00,  Effort = 3 | 2.02 | 0.67, 6.09 | .948 |
| $2.50 vs. $3.00,  Effort = 3 | 1.12 | 0.34, 3.73 | .999 |
| $0.50 vs. $1, Effort = 20 | 9.26 | 4.04, 21.24 | <.001 |
| $0.50 vs. $1.50, Effort = 20 | 10.34 | 4.53, 23.62 | <.001 |
| $0.50 vs. $2.00, Effort = 20 | 28.18 | 10.32, 76.96 | <.001 |
| $0.50 vs. $2.50, Effort = 20 | 31.12 | 11.51, 84.19 | <.001 |
| $0.50 vs. $3.00, Effort = 20 | 47.06 | 15.69, 141.22 | <.001 |
| $1.00 vs. $1.50, Effort = 20 | 1.12 | 0.48, 2.61 | .999 |
| $1.00 vs. $2.00, Effort = 20 | 3.04 | 1.10, 8.45 | .021 |
| $1.00 vs. $2.50, Effort = 20 | 3.36 | 1.22, 9.25 | .007 |
| $1.00 vs. $3.00,  Effort = 20 | 5.08 | 1.67, 15.48 | <.001 |
| $1.50 vs. $2.00,  Effort = 20 | 2.73 | 0.98, 7.54 | .058 |
| $1.50 vs. $2.50,  Effort = 20 | 3.01 | 1.10, 8.25 | .020 |
| $1.50 vs. $3.00,  Effort = 20 | 4.55 | 1.50, 13.81 | <.001 |
| $2.00 vs. $2.50, Effort = 20 | 1.11 | 0.35, 3.51 | .999 |
| $2.00 vs. $3.00,  Effort = 20 | 1.67 | 0.48, 5.79 | .999 |
| $2.50 vs. $3.00,  Effort = 20 | 1.51 | 0.44, 5.20 | .999 |
| $0.50 vs. $1, Effort = 37 | 3.76 | 1.71, 8.26 | <.001 |
| $0.50 vs. $1.50, Effort = 37 | 11.40 | 4.90, 26.54 | <.001 |
| $0.50 vs. $2.00, Effort = 37 | 19.78 | 8.15, 48.01 | <.001 |
| $0.50 vs. $2.50, Effort = 37 | 26.18 | 10.58, 64.82 | <.001 |
| $0.50 vs. $3.00, Effort = 37 | 49.22 | 16.97, 142.77 | <.001 |
| $1.00 vs. $1.50, Effort = 37 | 3.04 | 1.35, 6.82 | <.001 |
| $1.00 vs. $2.00, Effort = 37 | 5.27 | 2.25, 12.34 | <.001 |
| $1.00 vs. $2.50, Effort = 37 | 6.97 | 2.92, 16.66 | <.001 |
| $1.00 vs. $3.00,  Effort = 37 | 13.11 | 4.66, 36.89 | <.001 |
| $1.50 vs. $2.00,  Effort = 37 | 1.74 | 0.71, 4.26 | .999 |
| $1.50 vs. $2.50,  Effort = 37 | 2.30 | 0.92, 5.74 | .115 |
| $1.50 vs. $3.00,  Effort = 37 | 4.32 | 1.48, 12.61 | <.001 |
| $2.00 vs. $2.50, Effort = 37 | 1.32 | 0.51, 3.43 | .999 |
| $2.00 vs. $3.00,  Effort = 37 | 2.49 | 0.83, 7.48 | .227 |
| $2.50 vs. $3.00,  Effort = 37 | 1.88 | 0.62, 5.74 | .999 |
| $0.50 vs. $1, Effort = 54 | 2.99 | 1.28, 6.97 | .002 |
| $0.50 vs. $1.50, Effort = 54 | 8.45 | 3.67, 19.50 | <.001 |
| $0.50 vs. $2.00, Effort = 54 | 28.83 | 11.82, 70.33 | <.001 |
| $0.50 vs. $2.50, Effort = 54 | 52.85 | 20.49, 136.34 | <.001 |
| $0.50 vs. $3.00, Effort = 54 | 60.56 | 23.18, 158.22 | <.001 |
| $1.00 vs. $1.50, Effort = 54 | 2.83 | 1.31, 6.15 | .001 |
| $1.00 vs. $2.00, Effort = 54 | 9.66 | 4.20, 22.19 | <.001 |
| $1.00 vs. $2.50, Effort = 54 | 17.70 | 7.26, 43.15 | <.001 |
| $1.00 vs. $3.00,  Effort = 54 | 20.29 | 8.21, 50.11 | <.001 |
| $1.50 vs. $2.00,  Effort = 54 | 3.41 | 1.53, 7.62 | <.001 |
| $1.50 vs. $2.50,  Effort = 54 | 6.25 | 2.63, 14.84 | <.001 |
| $1.50 vs. $3.00,  Effort = 54 | 7.16 | 2.98, 17.24 | <.001 |
| $2.00 vs. $2.50, Effort = 54 | 1.83 | 0.74, 4.55 | .756 |
| $2.00 vs. $3.00,  Effort = 54 | 2.10 | 0.84, 5.28 | .272 |
| $2.50 vs. $3.00,  Effort = 54 | 1.15 | 0.43, 3.03 | .999 |
| $0.50 vs. $1.50, Effort = 70 | 3.19 | 1.39, 7.30 | <.001 |
| $0.50 vs. $2.00, Effort = 70 | 11.26 | 4.90, 25.85 | <.001 |
| $0.50 vs. $2.50, Effort = 70 | 23.00 | 9.60, 55.08 | <.001 |
| $0.50 vs. $3.00, Effort = 70 | 34.77 | 13.73, 88.07 | <.001 |
| $1.00 vs. $1.50, Effort = 70 | 1.44 | 0.66, 3.15 | .999 |
| $1.00 vs. $2.00, Effort = 70 | 5.07 | 2.32, 11.10 | <.001 |
| $1.00 vs. $2.50, Effort = 70 | 10.36 | 4.53, 23.69 | <.001 |
| $1.00 vs. $3.00,  Effort = 70 | 15.67 | 6.46, 37.99 | <.001 |
| $1.50 vs. $2.00,  Effort = 70 | 3.53 | 1.64, 7.60 | <.001 |
| $1.50 vs. $2.50,  Effort = 70 | 7.21 | 3.20, 16.23 | <.001 |
| $1.50 vs. $3.00,  Effort = 70 | 10.90 | 4.56, 26.05 | <.001 |
| $2.00 vs. $2.50, Effort = 70 | 2.04 | 0.92, 4.55 | .133 |
| $2.00 vs. $3.00,  Effort = 70 | 3.09 | 1.31, 7.30 | .002 |
| $2.50 vs. $3.00,  Effort = 70 | 1.51 | 0.62, 3.71 | .999 |

**Table** **S2** Post-hoc comparisons between effort levels by reward magnitude on choice data.

| Comparison | Odds Ratio | 95% CI | p (Bonferroni corrected) |
| --- | --- | --- | --- |
| 3 vs. 20,  Reward = $0.50 | 0.31 | 0.15, 0.66 | <.001 |
| 3 vs. 37,  Reward = $0.50 | 0.21 | 0.10, 0.45 | <.001 |
| 3 vs. 54,  Reward = $0.50 | 0.10 | 0.05, 0.22 | <.001 |
| 3 vs. 70,  Reward = $0.50 | 0.12 | 0.05, 0.26 | <.001 |
| 20 vs. 37,  Reward = $0.50 | 0.68 | 0.31, 1.46 | .999 |
| 20 vs. 54,  Reward = $0.50 | 0.32 | 0.14, 0.72 | <.001 |
| 20 vs. 70,  Reward = $0.50 | 0.37 | 0.17, 0.82 | .005 |
| 37 vs. 54,  Reward = $0.50 | 0.47 | 0.21, 1.07 | .104 |
| 37 vs. 70,  Reward = $0.50 | 0.55 | 0.24, 1.23 | .358 |
| 54 vs. 70,  Reward = $0.50 | 1.15 | 0.50, 2.67 | .999 |
| 3 vs. 20,  Reward = $1.00 | 0.76 | 0.33, 1.75 | .999 |
| 3 vs. 37,  Reward = $1.00 | 0.21 | 0.10, 0.46 | <.001 |
| 3 vs. 54,  Reward = $1.00 | 0.08 | 0.04, 0.18 | <.001 |
| 3 vs. 70,  Reward = $1.00 | 0.07 | 0.03, 0.15 | <.001 |
| 20 vs. 37,  Reward = $1.00 | 0.27 | 0.13, 0.60 | <.001 |
| 20 vs. 54,  Reward = $1.00 | 0.10 | 0.05, 0.23 | <.001 |
| 20 vs. 70,  Reward = $1.00 | 0.09 | 0.04, 0.20 | <.001 |
| 37 vs. 54,  Reward = $1.00 | 0.38 | 0.18, 0.79 | .002 |
| 37 vs. 70,  Reward = $1.00 | 0.32 | 0.15, 0.68 | <.001 |
| 54 vs. 70,  Reward = $1.00 | 0.86 | 0.40, 1.83 | .999 |
| 3 vs. 20,  Reward = $1.50 | 1.37 | 0.62, 3.02 | .999 |
| 3 vs. 37,  Reward = $1.50 | 1.02 | 0.46, 2.27 | .999 |
| 3 vs. 54,  Reward = $1.50 | 0.36 | 0.17, 0.76 | .001 |
| 3 vs. 70,  Reward = $1.50 | 0.16 | 0.07, 0.33 | <.001 |
| 20 vs. 37,  Reward = $1.50 | 0.75 | 0.33, 1.69 | .999 |
| 20 vs. 54,  Reward = $1.50 | 0.26 | 0.12, 0.57 | <.001 |
| 20 vs. 70,  Reward = $1.50 | 0.11 | 0.05, 0.25 | <.001 |
| 37 vs. 54,  Reward = $1.50 | 0.35 | 0.16, 0.76 | .001 |
| 37 vs. 70,  Reward = $1.50 | 0.15 | 0.07, 0.34 | <.001 |
| 54 vs. 70,  Reward = $1.50 | 0.44 | 0.21, 0.90 | .013 |
| 3 vs. 20,  Reward = $2.00 | 1.55 | 0.55, 4.35 | .999 |
| 3 vs. 37,  Reward = $2.00 | 0.74 | 0.29, 1.85 | .999 |
| 3 vs. 54,  Reward = $2.00 | 0.51 | 0.21, 1.23 | .320 |
| 3 vs. 70,  Reward = $2.00 | 0.23 | 0.10, 0.53 | <.001 |
| 20 vs. 37,  Reward = $2.00 | 0.48 | 0.17, 1.31 | .389 |
| 20 vs. 54,  Reward = $2.00 | 0.33 | 0.12, 0.87 | .014 |
| 20 vs. 70,  Reward = $2.00 | 0.15 | 0.06, 0.38 | <.001 |
| 37 vs. 54,  Reward = $2.00 | 0.69 | 0.29, 1.63 | .999 |
| 37 vs. 70,  Reward = $2.00 | 0.31 | 0.14, 0.70 | <.001 |
| 54 vs. 70,  Reward = $2.00 | 0.45 | 0.21, 0.98 | .038 |
| 3 vs. 20,  Reward = $2.50 | 0.95 | 0.31, 2.91 | .999 |
| 3 vs. 37,  Reward = $2.50 | 0.54 | 0.19, 1.53 | .983 |
| 3 vs. 54,  Reward = $2.50 | 0.52 | 0.18, 1.46 | .757 |
| 3 vs. 70,  Reward = $2.50 | 0.26 | 0.10, 0.70 | .001 |
| 20 vs. 37,  Reward = $2.50 | 0.57 | 0.21, 1.57 | .999 |
| 20 vs. 54,  Reward = $2.50 | 0.55 | 0.20, 1.50 | .931 |
| 20 vs. 70,  Reward = $2.50 | 0.27 | 0.10, 0.72 | .002 |
| 37 vs. 54,  Reward = $2.50 | 0.96 | 0.38, 2.41 | .999 |
| 37 vs. 70,  Reward = $2.50 | 0.48 | 0.20, 1.14 | .176 |
| 54 vs. 70,  Reward = $2.50 | 0.50 | 0.21, 1.19 | .249 |
| 3 vs. 20,  Reward = $3.00 | 1.29 | 0.38, 4.32 | .999 |
| 3 vs. 37,  Reward = $3.00 | 0.91 | 0.28, 2.95 | .999 |
| 3 vs. 54,  Reward = $3.00 | 0.53 | 0.18, 1.53 | .936 |
| 3 vs. 70,  Reward = $3.00 | 0.35 | 0.12, 1.00 | .05 |
| 20 vs. 37,  Reward = $3.00 | 0.71 | 0.21, 2.42 | .999 |
| 20 vs. 54,  Reward = $3.00 | 0.41 | 0.14, 1.26 | .261 |
| 20 vs. 70,  Reward = $3.00 | 0.27 | 0.09, 0.82 | .010 |
| 37 vs. 54,  Reward = $3.00 | 0.58 | 0.20, 1.71 | .999 |
| 37 vs. 70,  Reward = $3.00 | 0.39 | 0.13, 1.12 | .118 |
| 54 vs. 70,  Reward = $3.00 | 0.66 | 0.26, 1.67 | .999 |

**Table** **S3** Post-hoc comparisons between conditions by APOE4 carrier status on vigor.

| Comparison | Mean difference | 95% CI | p (Bonferroni corrected) |
| --- | --- | --- | --- |
| Self-oriented vs. Prosocial, APOE4 non-carriers | -0.01 | -0.05, 0.04 | .807 |
| Self-oriented vs. Prosocial, APOE4 carriers | -0.10 | -0.17, -0.03 | .008 |
| APOE4 non carrier vs. APOE4 carrier, Self-oriented | -0.10 | -0.29, 0.08 | .279 |
| APOE4 non carrier vs. APOE4 carrier, Prosocial | -0.20 | -0.38, -0.01 | .037 |

**Figure** **S1** Sigmoid model fit curves for each participant for the prosocial (Panel A) and self-oriented (Panel B) reward conditions.

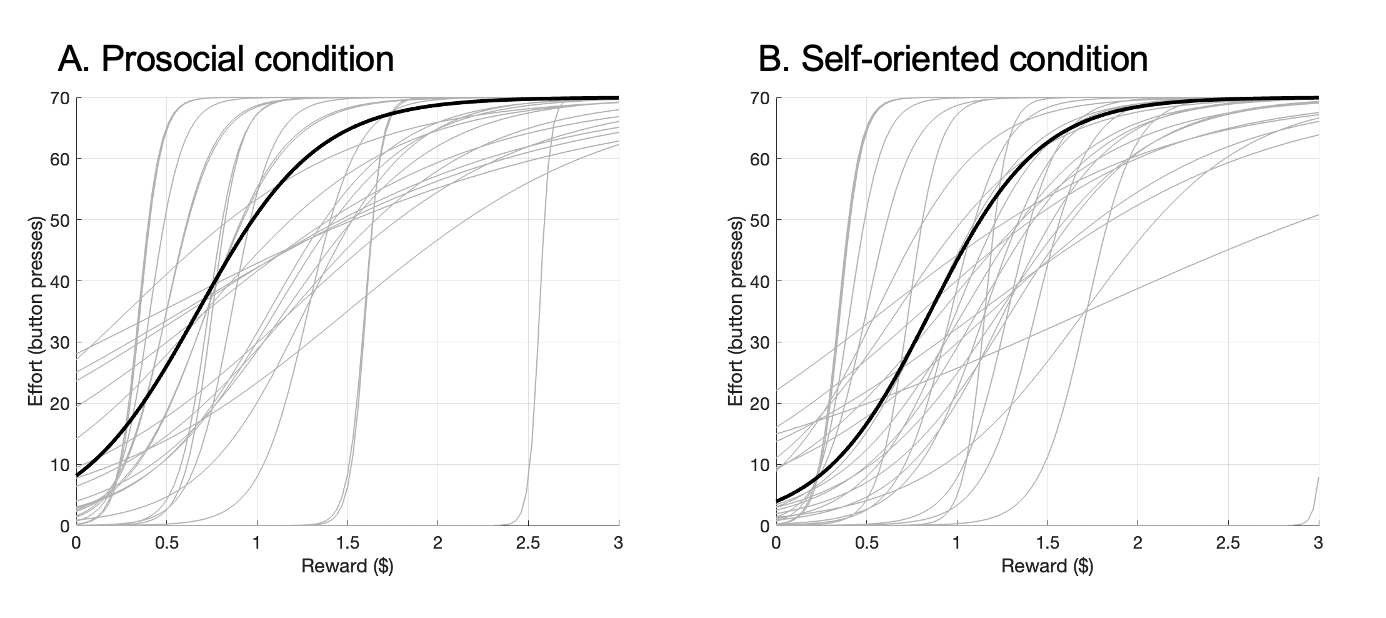

**Resting-State fMRI Results**

The vmPFC seed was primarily positively associated with regions within the default mode (e.g., medial prefrontal cortex, frontal poles, precuneus, hippocampus, parahippocampus), frontoparietal (e.g., dorsolateral prefrontal cortex), salience (e.g., anterior cingulate cortex, insula, thalamus) and limbic networks (orbitofrontal cortex, subcallosal cortex, nucleus accumbens, amygdala) and negatively associated with regions from the visual network (e.g., middle temporal gyrus, lateral occipital cortex, fusiform gyri), salience (supramarginal gyri), frontoparietal (e.g., inferior frontal gyri) and motor networks (primary motor cortex, supplementary motor cortex).

**Figure** **S2** Seed-to-voxel resting-state functional connectivity from the ventromedial prefrontal cortex seed.

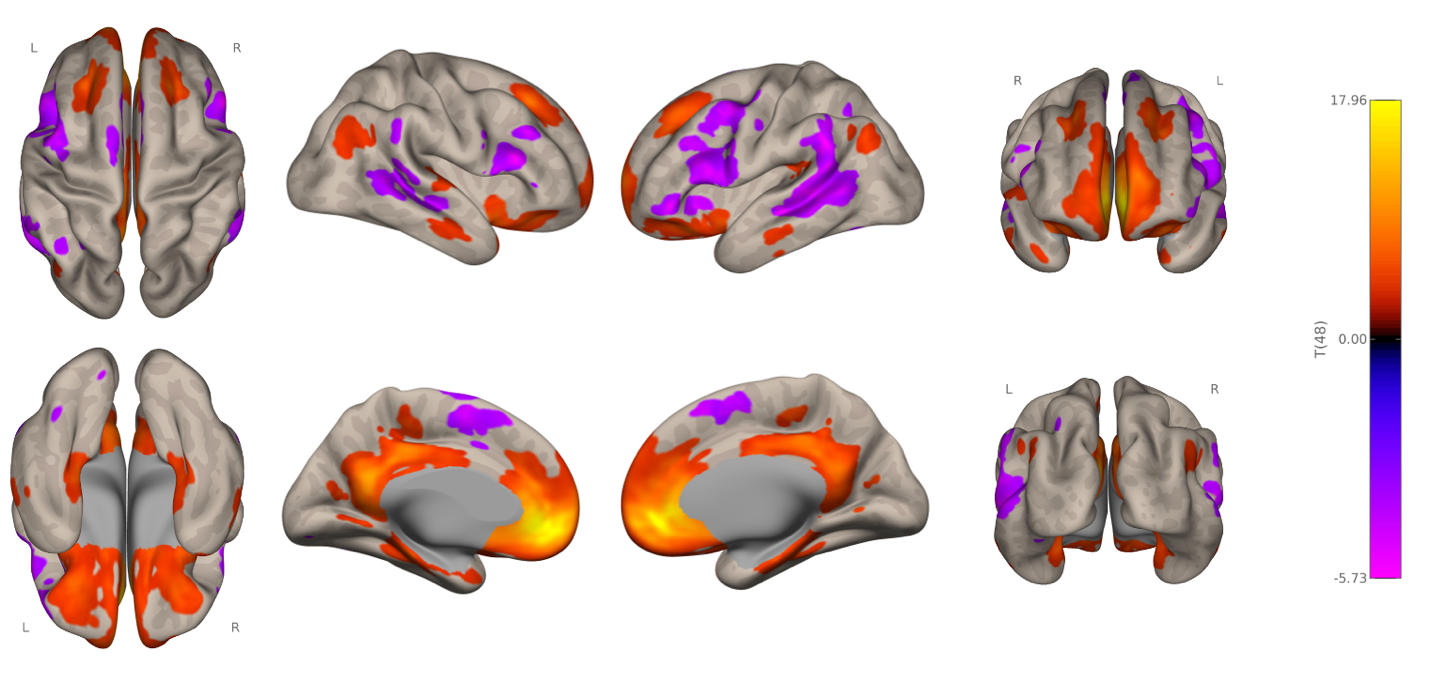

The ACC seed was positively associated with regions of the salience (insula, paracingulate cortex, thalamus), default (medial frontal cortex, posterior cingulate cortex, hippocampus, parahippocampal cortex, precuneus) and limbic networks (orbitofrontal cortex, temporal poles, subcallosal cortex, amygdala, nucleus accumbens) and negatively associated with regions from the visual network (lateral occipital cortex, temporo-occipital cortex, middle temporal gyri), frontoparietal network (inferior frontal gyri, dorsolateral prefrontal cortex), default mode network (frontal poles) and motor network (primary motor cortex).

**Figure** **S3** Seed-to-voxel resting-state functional connectivity from the anterior cingulate cortex seed.

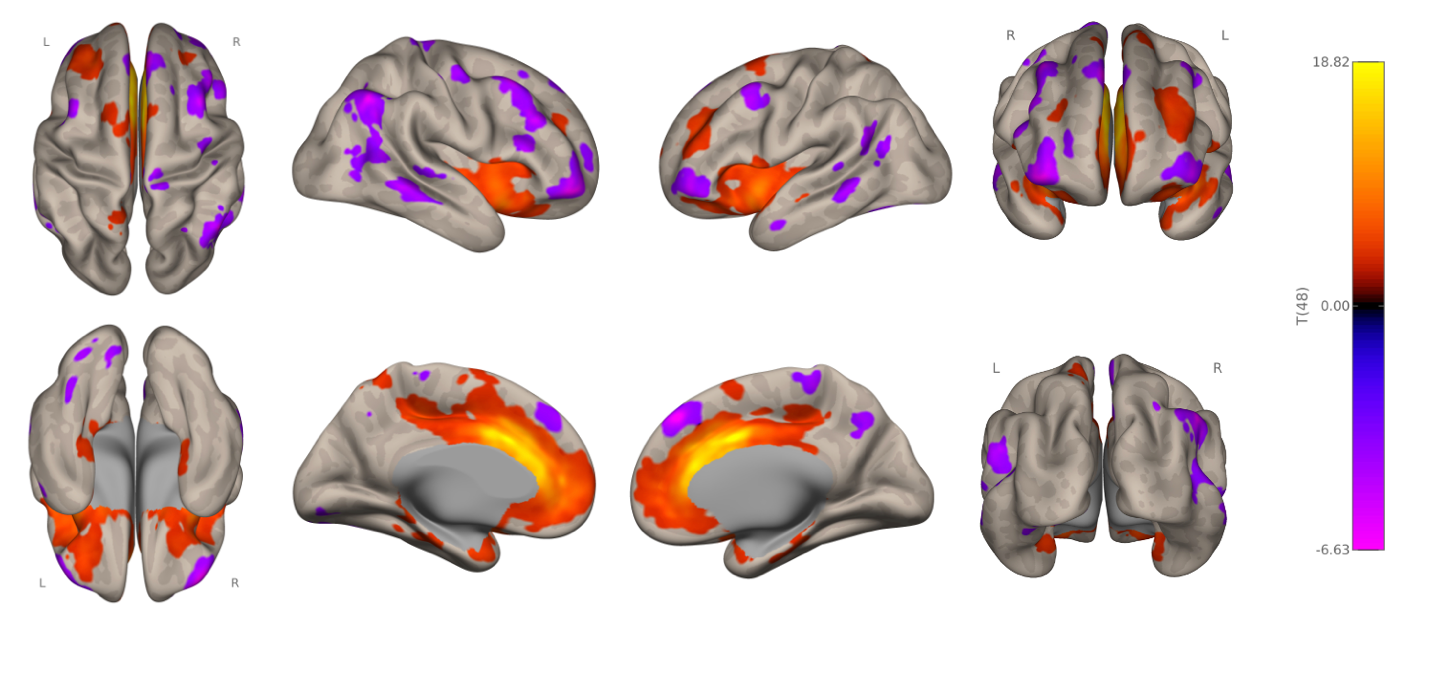

**Table** **S4** Beta estimates and 95% bootstrapped confidence intervals of robust regression results for seed-to-voxel analyses from the ventromedial prefrontal cortex and anterior cingulate cortex seeds.

| Connection | Beta | Bootstrapped 95% CI |
| --- | --- | --- |
| vmPFC-dmPFC | 0.40 | 0.17, 0.75 |
| ACC-IFG1 | 0.81 | 0.17, 1.49 |
| ACC_IFG2 | 0.41 | 0.02, 1.02 |
| ACC-rLOC | -0.81 | -1.50, -0.22 |
| ACC_rSPL | -0.57 | -1.28, -0.11 |
| ACC_rSTG | -1.08 | -1.62, -0.21 |
| ACC_rdlPFC | -0.79 | -1.64, -0.29 |

*Note.* vmPFC = ventromedial prefrontal cortex; ACC = anterior cingulate cortex; IFG1 = cluster 1 of inferior frontal gyrus; IFG2 = cluster 2 of inferior frontal gyrus; SPL = superior parietal lobule; m/sTG = middle/superior temporal gyrus; dlPFC = dorsolateral prefrontal cortex; LOC = lateral occipital cortex.

**Figure S4** Scatterplots depicting relationship between effort bias difference scores (self-oriented – prosocial) and seed-to-voxel rsFC to the ventromedial prefrontal cortex and anterior cingulate cortex seeds.

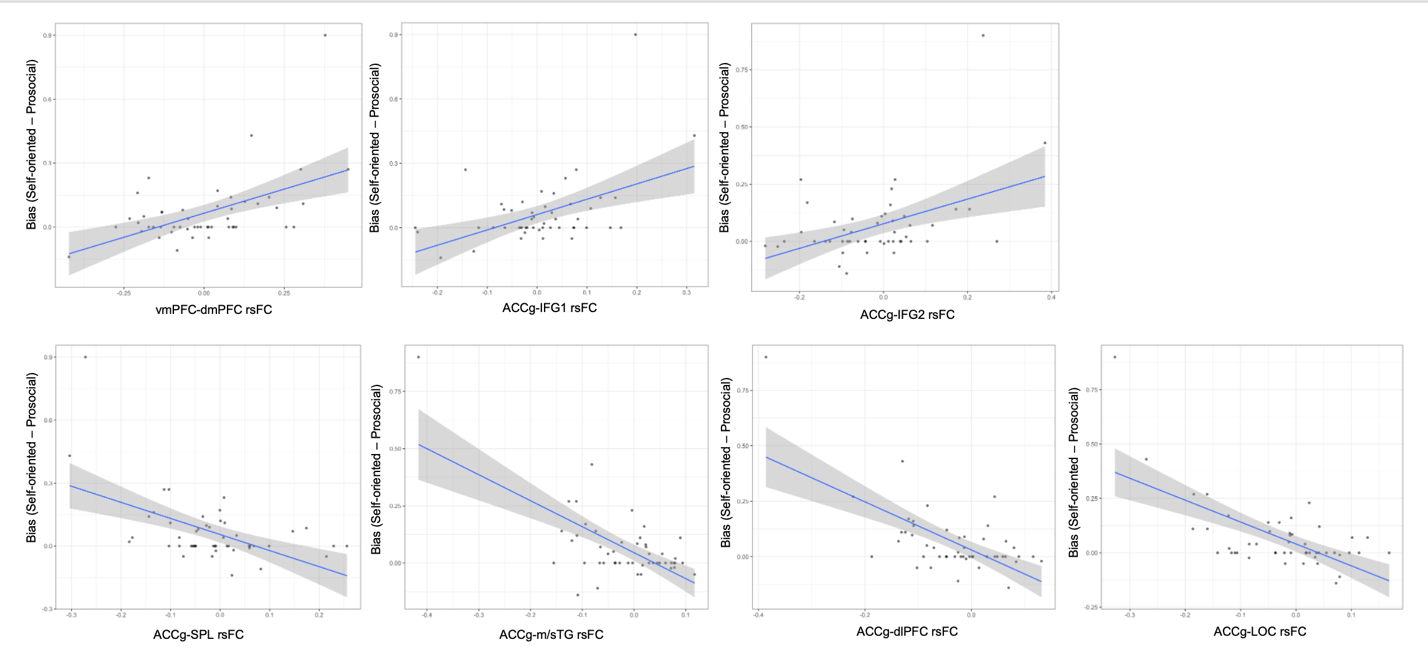

*Note.* vmPFC = ventromedial prefrontal cortex; ACC = anterior cingulate cortex; IFG1 = cluster 1 of inferior frontal gyrus; IFG2 = cluster 2 of inferior frontal gyrus; SPL = superior parietal lobule; m/sTG = middle/superior temporal gyrus; dlPFC = dorsolateral prefrontal cortex; LOC = lateral occipital cortex.

**Table** **S5** Beta estimates and 95% bootstrapped confidence intervals of robust regression results for post-hoc seed-to-voxel analyses from anterior and posterior ventromedial prefrontal cortex seeds derived from fc-MVPA.

| Connection | Beta | Bootstrapped 95% CI |
| --- | --- | --- |
| pvmPFC-rLOC | -1.53 | -1.91, -0.24 |
| pvmPFC-rdlPFC | 0.89 | 0.19, 1.79 |
| avmPFC-rTP | 0.90 | 0.30, 1.71 |
| avmPFC-lTP | 0.50 | 0.01, 1.20 |
| avmPFC-lLOC | -0.83 | -1.66, -0.46 |
| avmPFC-lpostcentral gyrus | -1.01 | -1.82, -0.39 |
| avmPFC-rFP | 0.59 | 0.25, 1.01 |
| avmPFC-lTPJ | -0.84 | -1.62, -0.13 |

*Note.* pvmPFC = posterior ventromedial prefrontal cortex seed (cluster 1 in manuscript); avmPFC = anterior ventromedial prefrontal cortex seed (cluster 3 in manuscript); rLOC = right lateral occipital cortex; rdlPFC = right dorsolateral prefrontal cortex; rTP = right temporal pole; lTP = left temporal pole; lLOC = left lateral occipital cortex; rFP = right frontal pole; lTPJ = left temporoparietal junction.

**Figure S5** Scatterplots depicting relationship between effort bias difference scores (self-oriented – prosocial) and post-hoc seed-to-voxel rsFC to the anterior ventromedial prefrontal cortex and posterior ventromedial prefrontal cortex seeds identified from fc-MVPA.

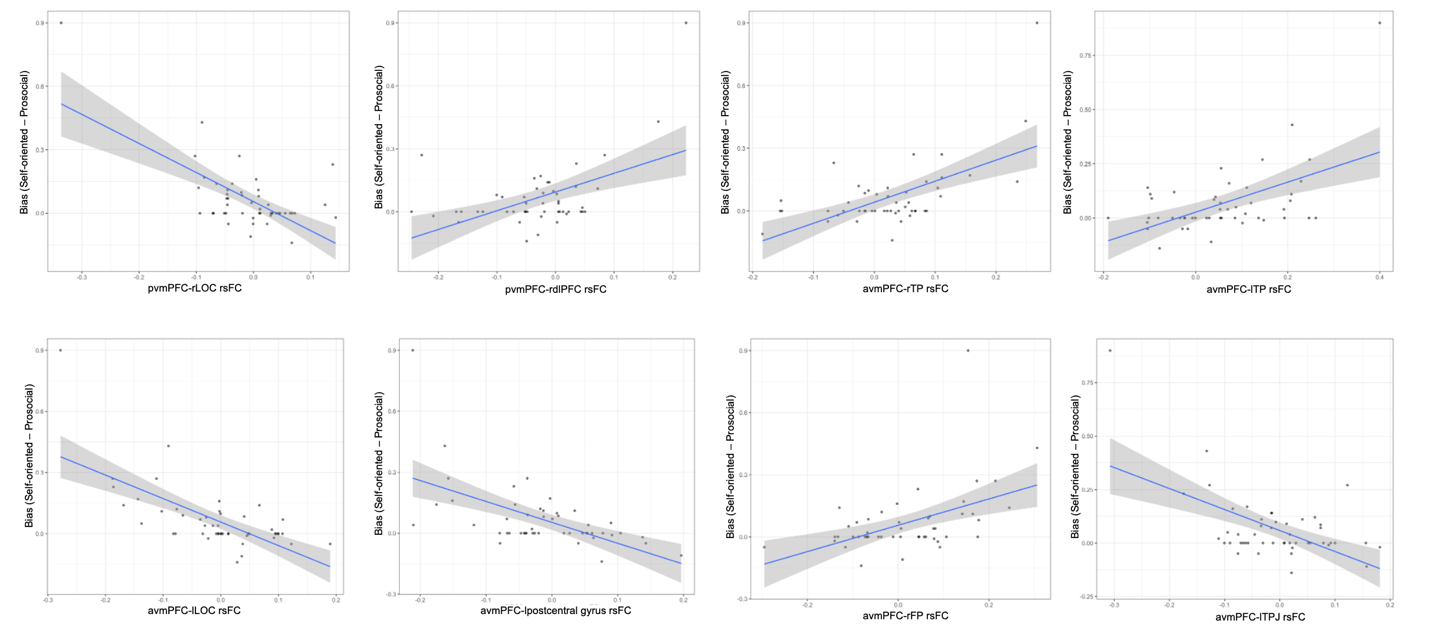

*Note.* pvmPFC = posterior ventromedial prefrontal cortex seed (cluster 1 in manuscript); avmPFC = anterior ventromedial prefrontal cortex seed (cluster 3 in manuscript); rLOC = right lateral occipital cortex; rdlPFC = right dorsolateral prefrontal cortex; rTP = right temporal pole; lTP = left temporal pole; lLOC = left lateral occipital cortex; rFP = right frontal pole; lTPJ = left temporoparietal junction.

A robust regression analysis confirmed that the negative association between dACC-rNAcc rsFC and effort bias (averaged across both conditions) held after accounting for outliers, β = -0.41, 95% CI [-0.88, -0.08].

**Figure** **S6** Scatterplot depicting the negative association between dorsal anterior cingulate cortex (dACC) – right nucleus accumbens (rNAcc) resting-state functional connectivity (rsFC) and bias away from effort across both conditions.

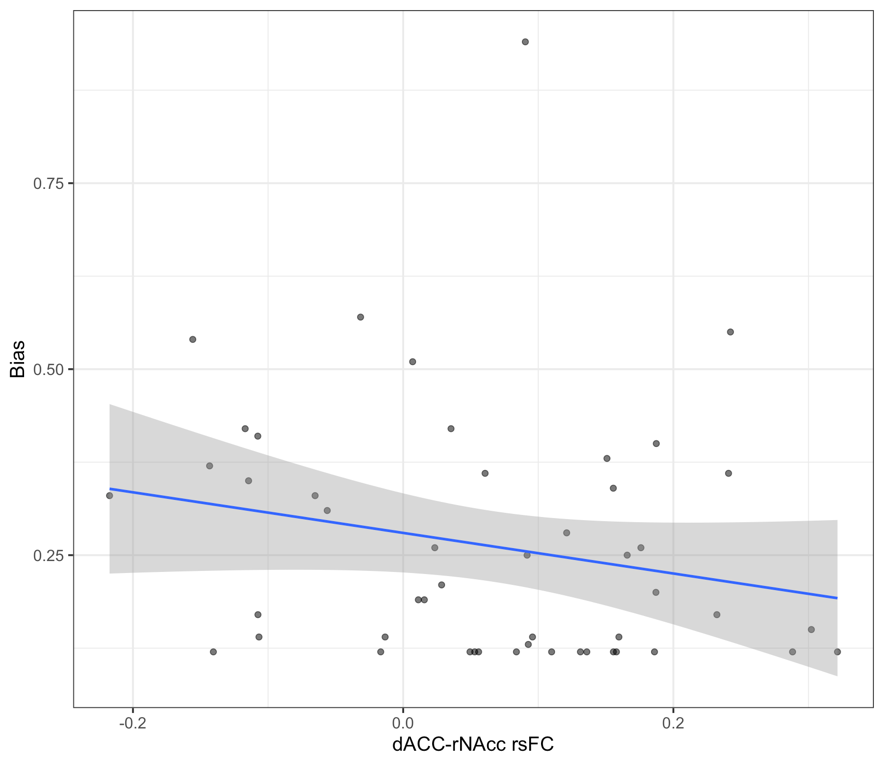
